# Development of a future prediabetes risk assessment model for individuals with normal glucose levels using efficiency scores obtained from data envelopment analysis

**DOI:** 10.1101/2025.05.21.25328057

**Authors:** Sho Nakamura, Rina Inoue, Hiroto Narimatsu

**Affiliations:** Graduate School of Health Innovation, Kanagawa University of Human Services, Kawasaki, Kanagawa, Japan; Cancer Prevention and Control Division, Kanagawa Cancer Center Research Institute, Yokohama, Kanagawa, Japan; CIKOP, specified non-profit organization, Yamagata, Yamagata, Japan; Department of Medical Genetics, Kanagawa Cancer Center, Yokohama, Kanagawa, Japan

## Abstract

**Background:** Identifying healthy individuals at risk of prediabetes for primary prevention is crucial, as current tools often focus on secondary prevention. We investigated whether efficiency scores, derived from data envelopment analysis (DEA), predict prediabetes development in a healthy population.

**Methods:** This historical cohort study analyzed annual health checkup data. Cox proportional hazards analysis assessed the relationship between efficiency scores and incident prediabetes. A classification tree analysis was also performed, incorporating efficiency scores, hemoglobin A1c (HbA1c), and other diabetes-related variables.

**Results:** The cohort comprised 923 individuals (49.7% female), with a mean efficiency score of 0.72 (0.07). During follow-up, 175 participants developed prediabetes (79.3 per 1,000 person-years). A 0.1-point increase in efficiency score was associated with an adjusted hazard ratio of 0.51 (95% CI 0.39-0.68, p < 0.0001) for prediabetes, while a 0.1% increase in HbA1c yielded an adjusted hazard ratio of 2.26 (95% CI 1.88-2.71, p < 0.0001). The classification tree identified a high-risk group of 31 individuals (3.4%) with 12.1% sensitivity and 98.7% specificity.

**Discussion:** Efficiency scores are linked to the 3-year risk of prediabetes in healthy subjects. The combined use of DEA and classification tree analysis presents a potentially valuable approach for primary prevention strategies in clinical practice.

## Introduction

Diabetes mellitus is a global health concern, affecting over 500 million individuals worldwide. With approximately 10% of adults aged 20-79 years having diabetes and about half remaining undiagnosed, the need for effective prevention strategies is critical (1, 2). Current interventions for type 2 diabetes mellitus (hereafter referred to as “diabetes”), which accounts for 90% of diabetes cases, primarily focus on secondary prevention through early detection and treatment (3, 4).

Standard screening methods include fasting plasma glucose (FPG) and glycated hemoglobin A1c (HbA1c) tests. However, these tests may not capture all cases of glucose intolerance, as some individuals with normal FPG and HbA1c levels may still meet diabetes criteria when subjected to an oral glucose tolerance test (OGTT) (1, 5, 6).

While various risk assessment tools exist for identifying prediabetes and predicting future diabetes incidence in general populations (7–13), a significant gap remains in the literature: no previous study has aimed to predict prediabetes among healthy individuals who screened negative according to FPG and/or HbA1c levels. Among individuals diagnosed with normal glucose tolerance, as evaluated by FPG, HbA1c, and OGTT, 11.5 and 2.3 per 1,000 person-years developed diabetes and prediabetes, respectively (14). Predicting prediabetes among individuals with normal screening results is essential for the primary prevention of diabetes development. We hypothesized that data envelopment analysis (DEA), a non-parametric technique, could offer a novel approach to predict prediabetes among individuals with normal screening results. DEA, originating from operations research and commonly used in benchmarking, calculates an “efficiency score” for each decision-making unit (DMU) by comparing its inputs and outputs relative to its peers. In this study, we conceptualize “efficiency” as an individual’s ability to maintain a favorable health output (i.e., lower HbA1c, indicating better glucose homeostasis) given a set of health-related inputs (e.g., BMI, blood pressure). A lower efficiency score might reflect a less optimal interplay of various (measured or unmeasured) factors influencing glucose metabolism, even if individual inputs are within normal ranges. Our previous studies suggests that such DEA-derived efficiency score can reveal risk gradations for hypertension and dyslipidemia within populations initially deemed healthy by standard tests (15, 16). These studies demonstrated that efficiency scores may thus identify “at risk of risks” who could benefit from early preventive attention and encourage them to adopt healthier lifestyles (17). The present study aimed to explore the utility of DEA-derived efficiency scores as a complementary tool to predict the development of prediabetes specifically in individuals with normal baseline HbA1c levels. We posited that this approach, by holistically assessing multiple factors, might capture subtle inefficiencies in health maintenance not apparent through individual risk factor assessment alone. Furthermore, we combined the efficiency score with classification and regression tree (CART) analysis to identify an ideal population for primary prevention interventions (18).

Current guidelines recommend re-evaluating glucose tolerance at three-year intervals for individuals with normal screening results (1). If we can identify the population “at risk of risks” for developing prediabetes within 3 years, among those whose FPG or HbA1c are within the normal range, it would be possible to provide them further assessment by OGTT or prioritize them for intensive primary preventive intervention.

We examined whether the efficiency score obtained from DEA was associated with the development of prediabetes using the health checkup data of residents of a city in Japan. We also evaluated whether we could identify the specific population to be prioritized for intervention in clinical practice using decision tree analysis.

## Methods

### Data acquisition

This study used annual health checkup data from residents of Kaminoyama City, Japan, whose insurance is covered by the National Health Insurance Program, part of the universal health insurance system in Japan. Approximately one-third of the population is covered by national health insurance (19). The annual health checkup for the population is known specifically as “tokutei-kenkou-shinsa” (specific health checkup) and is stipulated by the Assurance of Medical Care for Elderly People Act (5). We obtained health checkup data from 2008 to 2013 from Kaminoyama City. The health checkups and the universal health insurance system in Japan have been described in detail elsewhere (5).

### Study design

We conducted a historical cohort study. Figure 1 presents a flow diagram of study participants. The inclusion criteria for this study were individuals with HbA1c < 5.6% and age < 74 years at baseline. These criteria ensure that our study population consisted of individuals with normal HbA1c levels who were eligible for primary prevention strategies, whom we refer to as healthy individuals hereafter. The inclusion date was the specific health checkup date, and observations were censored at the first time outcomes developed or at the end of the observation period. The end of the observation was the specific health checkup date 3, 2, and 1 fiscal year later for participants included between 2008 and 2010 and for 2011 and 2012. We chose a maximum 3-year follow-up period as it aligns with current guidelines recommending re-evaluation of glucose tolerance at three-year intervals for individuals with normal screening results (1). Participants ≥ 74 years of age in each year were excluded because they would be shifted to late-stage elderly insurance from the next year (5, 20), making them no longer eligible for specific health checkups and, therefore, unable to be followed. This study was approved by the Ethics Committee of Kanagawa Cancer Center (Yokohama, Japan; approval no. 27KEN-73). As this research involves the secondary use of anonymized data, informed consent was not obtained

**Figure 1.**
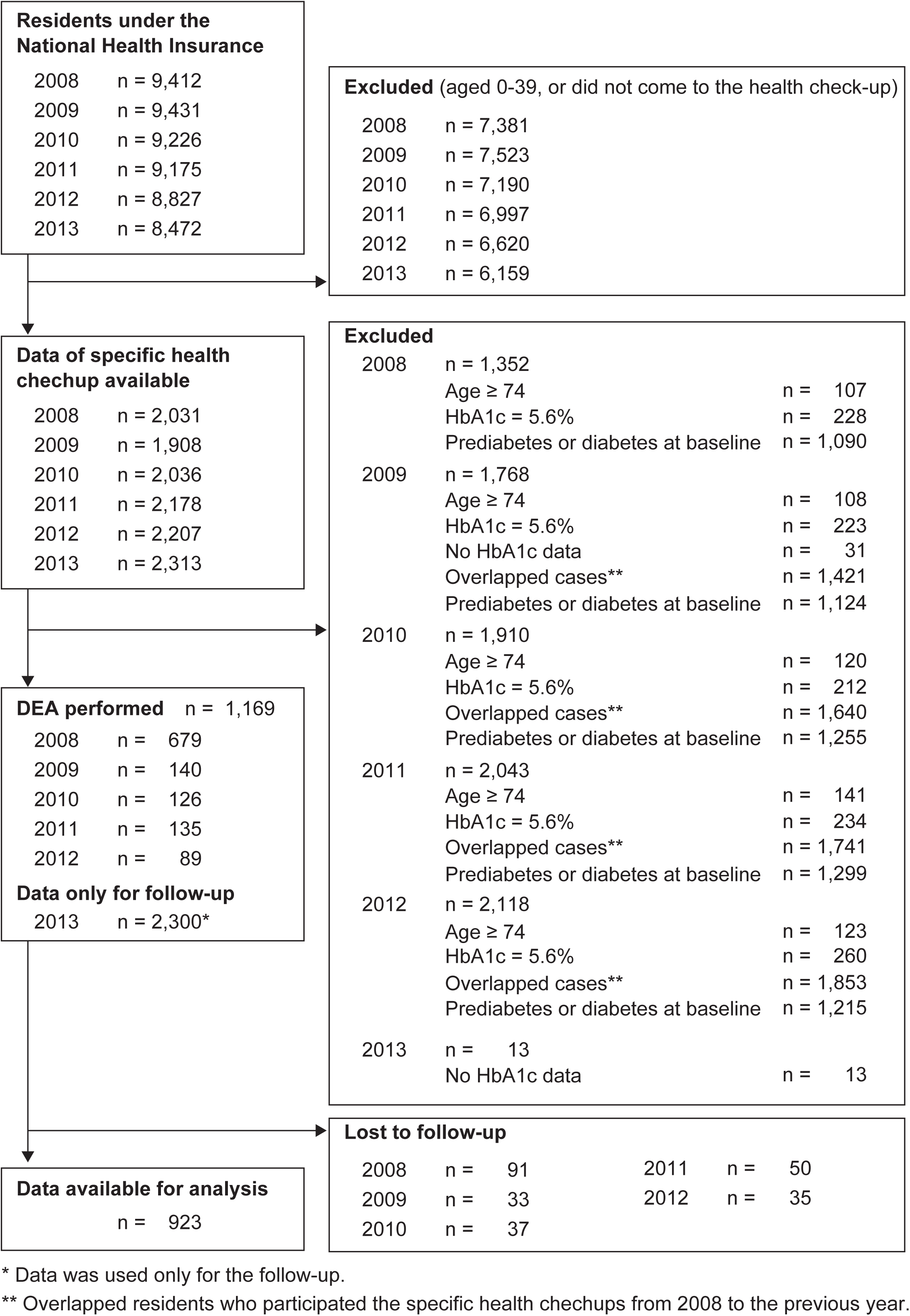
Flow diagram of the dataset.

### Variables in the analysis

We used data obtained from specific health checkups (5). The accuracy of the examination results was guaranteed in accordance with the standards of the Ministry of Health, Labor, and Welfare of Japan. We converted the HbA1c value to an equivalent value in accordance with the National Glycohemoglobin Standardization Program by adding 0.4% to the value from the Japan Diabetes Society (21). We defined prediabetes as HbA1c values of 5.7%, including diabetes (HbA1c ≥ 6.5%), and set this as an outcome (1). Participants with prediabetes at baseline and those taking antihyperglycemic medication were excluded. Participants with an HbA1c value of 5.6% in each year were also excluded to reduce bias because the treatment guidelines for diabetes, published by the Japanese Diabetes Society, recommend that these subjects undergo an OGTT (21, 22). Missing values for hemoglobin were replaced with mean values according to sex; more specifically, 14.8 g/dl for males (n = 4) and 13.0 g/dl for females (n = 9). Daily alcohol intake was evaluated by multiplying the frequency (rarely, 0; occasionally, 0.5; and every day, 1) and the amount (< 20; ≥ 20 to < 40; ≥ 40 to < 60; and ≥ 60 g) of alcohol intake per day as 0.5, 1.5, 2.5, and 3.5, respectively) and was treated as a continuous variable (units/day).

### DEA analysis

DEA was performed to calculate the efficiency score, which was hypothesized to serve as the risk score of the DMUs—more specifically, each participant. The output-oriented continuous returns-to-scale Charnes-Cooper-Rhodes DEA model was used to calculate the efficiency score (23). DMUs are responsible for obtaining the results (outputs) produced by the resources (inputs), and no a priori function is required to calculate the relative efficiency in DEA (23, 24). In an output-oriented model, the DMU that yields the maximum output for a given input is the most efficient. Selected inputs were based on known risk factors used in the risk prediction tools for type 2 diabetes (1, 8–11): BMI, waist circumference (WC), systolic and diastolic blood pressure, gamma-glutamyl transferase, triglyceride, and high-density lipoprotein cholesterol. The output was HbA1c level. The output-oriented approach was selected because it aligns with the clinical objective of maximizing health outcomes, minimizing HbA1c as an undesirable output, for a given set of inputs. In this context, the inefficiency evaluated in this model reflects a DMU’s relative sub-optimality in achieving a low HbA1c compared to peers with similar input profiles, potentially due to unmeasured factors like genetic predispositions, lifestyle differences, variations in metabolic resilience or the built environment. Therefore, BMI, WC, blood pressure, gamma-glutamyl transferase, triglyceride, and HbA1c levels were reversed by subtracting the sum of the maximum and minimum values of each variable. While DEA considers multiple inputs and outputs, the benchmarking result is presented as a single value: the efficiency score. The “dea” function of the Benchmarking package was used to calculate the inefficiency score (25). The reciprocal value of the calculated inefficiency score was used as the efficiency score from 0 to 1, with higher scores indicating higher efficiency. Another model was also constructed, restricting inputs, such as BMI, WC, and blood pressure, to factors that could be obtained without laboratory investigations. The efficiency score obtained in this model is referred to as the efficiency score (simple). The efficiency score (simple) assumes a different situation for clinical use; for example, in the healthcare setting, where only HbA1c test is available (e.g., by point-of-care testing) and results from other laboratory investigations are unavailable.

### Statistical analysis

Statistical analyses were performed using R version 4.2.3 (26). Pearson’s product-moment correlation between HbA1c level and efficiency scores was calculated. The unadjusted time to the development of prediabetes was determined using Kaplan–Meier analysis. Cox proportional hazards methods were used to estimate hazard ratio (HR) and corresponding 95% confidence interval (CI) of developing outcomes using the “coxph” function in the survival package. (27) Three models for efficiency score (simple), efficiency score, and HbA1c level were constructed. Each model was adjusted for the following variables to avoid severe bias caused by omitting variables that influence the outcome: age; sex; medication status for hypertension; medication status for dyslipidemia; smoking status; alcohol intake; and blood test results. Blood test parameters included low-density lipoprotein cholesterol (LDL-c), aspartate aminotransferase (AST), alanine transaminase (ALT), and hemoglobin. Because the efficiency score (simple) assumes a situation in which blood test results are unavailable, blood test results were excluded from the efficiency score (simple) model. The linearity assumption was checked by assessing the functional form of a continuous variable by plotting Martingale residuals using the “ggcox” function in the Survminer package (28). Any variable that could not achieve linearity was categorized as a continuous variable. Alcohol intake was categorized into 2 groups (< 1 and ≥ 1 units/day), as were LDL-c (< 130 and ≥ 130 mg/dl), AST (< 23 and ≥ 23 U/l), and ALT (< 19 and ≥ 20 U/l), while hemoglobin was categorized into 3 groups (< 14, ≥ 14 to ≤ 18, and > 18 g/l for males; and < 12, ≥ 12 to ≤ 16, and > 16 g/l for females). The multicollinearity of each adjusted model was checked through linear regression using the “vif” function of the DAAG package. The proportional hazards assumption was checked based on scaled Schoenfeld residuals using the “cox.zph” function in the survival package. For all variables, including each of the covariates, the global test demonstrated that the assumption of proportional hazards appeared to be supported. Influential observations were tested for by dfbeta residuals using the “ggcox” diagnostics function in the survminer package with a threshold of 0.066 = 2/√923 (28, 29). None of the observations exceeded this threshold.

In addition, decision tree models were applied to predict the development of prediabetes, in accordance with the Transparent Reporting of a multivariable prediction model for Individual Prognosis or Diagnosis (TRIPOD) reporting guidelines (30). The decision tree analysis, known as “CART,” is useful for clinical decision-making owing to its advantages such as ease of interpretation (31, 32). The CART algorithm in the “rpart” function of the rpart package was used. The splitting criterion employed by the rpart algorithm for classification trees is the Gini impurity index, which measures the extent to which any item from the node would be incorrectly classified if it were randomly labeled according to the distribution of labels in the node. The minimum terminal node size was set to 20 observations. The optimal cost-complexity pruning (CP) parameters were obtained by CP from the model with the largest area under the receiver operating characteristic curve (AUC) by five-fold cross-validation with 10 times repeat using the “train” function in the caret package. Two models were constructed for CART analysis, and the variables used in the adjusted Cox proportional hazards models for efficiency scores with HbA1c were used as inputs to the decision tree model to predict the future development of outcomes. Continuous variables were treated as such in the CART analysis. Additionally, we conducted a decision curve analysis using net benefit to assess the clinical utility of our model, and compared the performance of our model against the strategies of ‘treat all’ and ‘treat none’, evaluating the net benefit across different threshold probabilities (33).

## Results

Add your results here. Among the 1169 participants who underwent DEA, data from 923 (79.0%) were available for analysis (Figure 1). Dropouts accounted for 16.6% (153/923) of censoring. Baseline characteristics of the patients are summarized in Table 1. The mean (standard deviation) of the efficiency score and efficiency score (simple) was 0.72 (0.07), and 0.79 (0.07), respectively. The correlation between HbA1c level and efficiency scores were r(921) = −0.37 (95% confidence interval [CI] −0.42 to −0.31; P < 0.001) for efficiency score, and r(921) = −0.32 (95% CI −0.38 to −0.27; P < 0.001) for efficiency score (simple).

**Table 1.** Baseline characteristics of the analyzed residents.

| Variables | Mean (SD) |
| --- | --- |
| Toal subjects | n = 923 |
| Age (years) | 59.9 (8.9) |
| Sex (women), n (%) | 459 (49.7) |
| BMI (kg/m<sup>2</sup>) | 22.7 (2.9) |
| Waist circumference (cm) | 82.0 (8.3) |
| Blood pressure, systolic (mmHg) | 130.4 (19.6) |
| Blood pressure, diastolic (mmHg) | 78.8 (11.8) |
| Triglyceride (md/dl) | 103.9 (67.9) |
| HDL cholesterol (md/dl) | 67.9 (17.6) |
| LDL cholesterol (md/dl) | 121.3 (28.7) |
| AST (U/l) | 24.1 (12.6) |
| ALT (U/l) | 22.7 (14.1) |
| GGT (IU/l) | 36.7 (49.6) |
| Hemoglobin (g/dl)<sup>†</sup> | 13.9 (1.3) |
| HbA<sub>1c</sub> (median [IQR]) | 5.4 (5.3, 5.5) |
| Specific health guidance category (n [%])<sup>‡</sup> |  |
| Information provision | 783 (84.8) |
| Motivational health guidance | 113 (12.2) |
| Intensive health guidance | 27 (2.9) |
| On treatment for hypertension (n [%]) | 718 (77.8) |
| On treatment for dyslipidemia (n [%]) | 852 (92.3) |
| Smoking status (current), n (%) | 176 (19.1) |
| Daily alcohol intake (unit [IQR])<sup>§</sup> | 0.3 [0.0, 0.8] |
| Efficiency score (simple) | 0.72 (0.07) |
| Efficiency score (clinical) | 0.79 (0.07) |

Data are shown as mean (standard deviation) unless otherwise specified.
BMI, body mass index; HDL, high density lipoprotein; LDL, low density lipoprotein; AST, aspartate aminotransferase; ALT, alanine aminotransferase; GGT, gamma-glutamyl transpeptidase; HbA<sub>1c</sub>, glycated hemoglobin A<sub>1c</sub>; IQR, inter-quartile range.
<sup>†</sup> Missing value was replaced with mean of each sex; 14.8 g/dl for men (n = 4) and 13.0 g/dl for women (n = 9).
<sup>‡</sup> Refer to the publication by the OECD for the details (reference number 5).
<sup>§</sup> One unit corresponds to 20 g of alcohol.

The cumulative incidence curve derived from Kaplan–Meier analysis is presented in Figure 2. A total of 175 participants developed outcomes, with an incidence rate of 79.3 per 1,000 person-years. Only one participant exceeded HbA1c by 6.5%. The results of the Cox proportional hazards model are summarized in Table 2. Adjusted HRs observed for a 0.1-point increase in efficiency score (simple), efficiency score, and 0.1% increase in HbA1c were 0.56 (95% CI 0.43–0.73; P < 0.001), 0.51 (95% CI 0.39–0.68; P < 0.001), and 2.26 (95% CI 1.88–2.71; P < 0.001), respectively.

**Figure 2.**
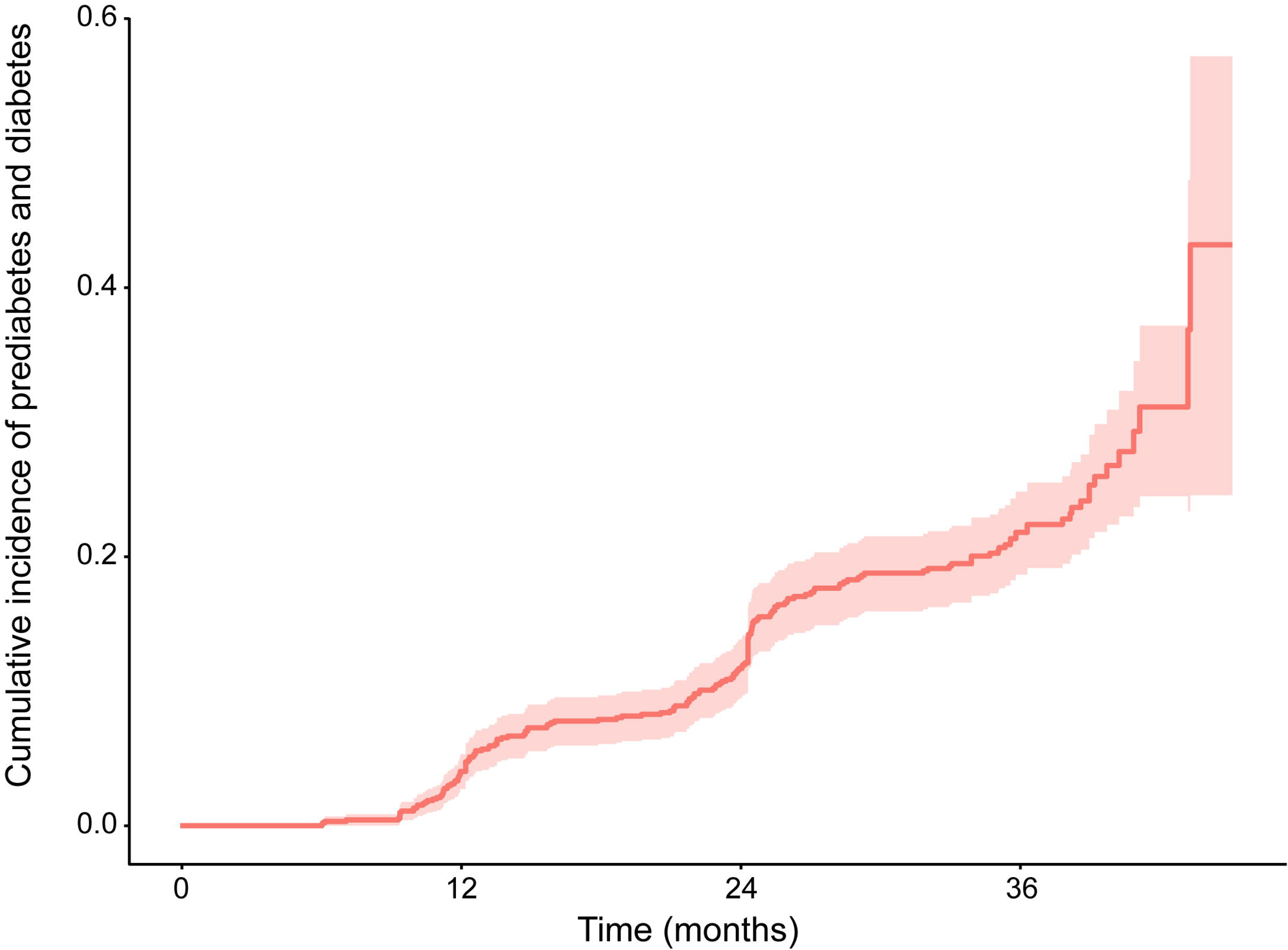
Cumulative incidence curve of development prediabetes and diabetes. Pale red area shows the 95% confidence interval.

**Table 2.**
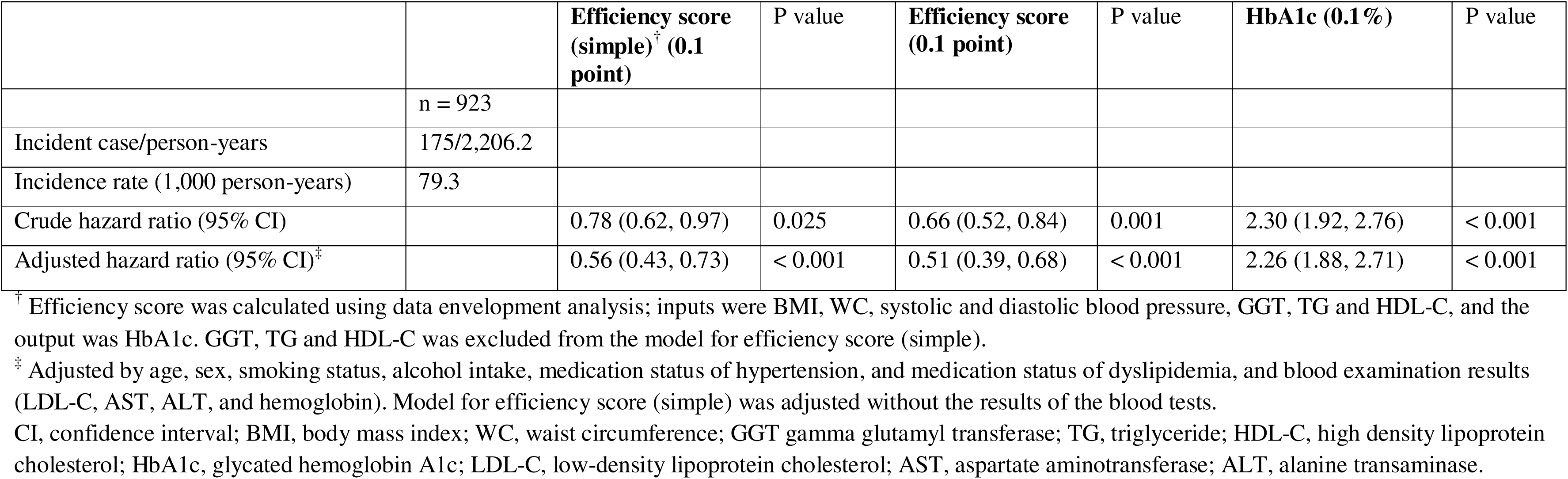
Hazard ratios for development of prediabetes.

The results obtained from the CART models are presented in Figure 3. The CP parameters for the simple efficiency score (simple) model were 5.71×103, and 11.4×103 for efficiency score model. From the simple efficiency score (simple), two terminal nodes were classified as positive (i.e., likely to develop prediabetes). In one of these terminal nodes, 17/26 (65.4%) participants developed outcomes: HbA1c 5.5%; efficiency score (simple) < 0.74; and age ≥ 70 years. In the other terminal node, 14/24 (58.3%) participants developed outcomes: HbA1c 5.5%; efficiency score (simple) < 0.74; age < 70 years; and taking medication for hypertension. For the efficiency score model, the terminal node with participants whose HbA1c level was 5.5%, efficiency score (simple) < 0.78, age ≥ 64, and LDL-c < 120 mg/dl were classified as positive. The model performance measures are shown in Figure 3. The efficiency score model demonstrated an overall sensitivity of 12.1%, a specificity of 98.7%, a positive likelihood ratio of 9.0, a negative likelihood ratio of 0.9. Figure 4 presents the results of the decision curve analysis for efficiency score model, illustrating the change in net benefit across threshold probabilities. The model demonstrated higher net benefit compared to other strategies (treat all, treat none) over a wide range of threshold probabilities from approximately 0.05 to 0.6.

**Figure 3.**
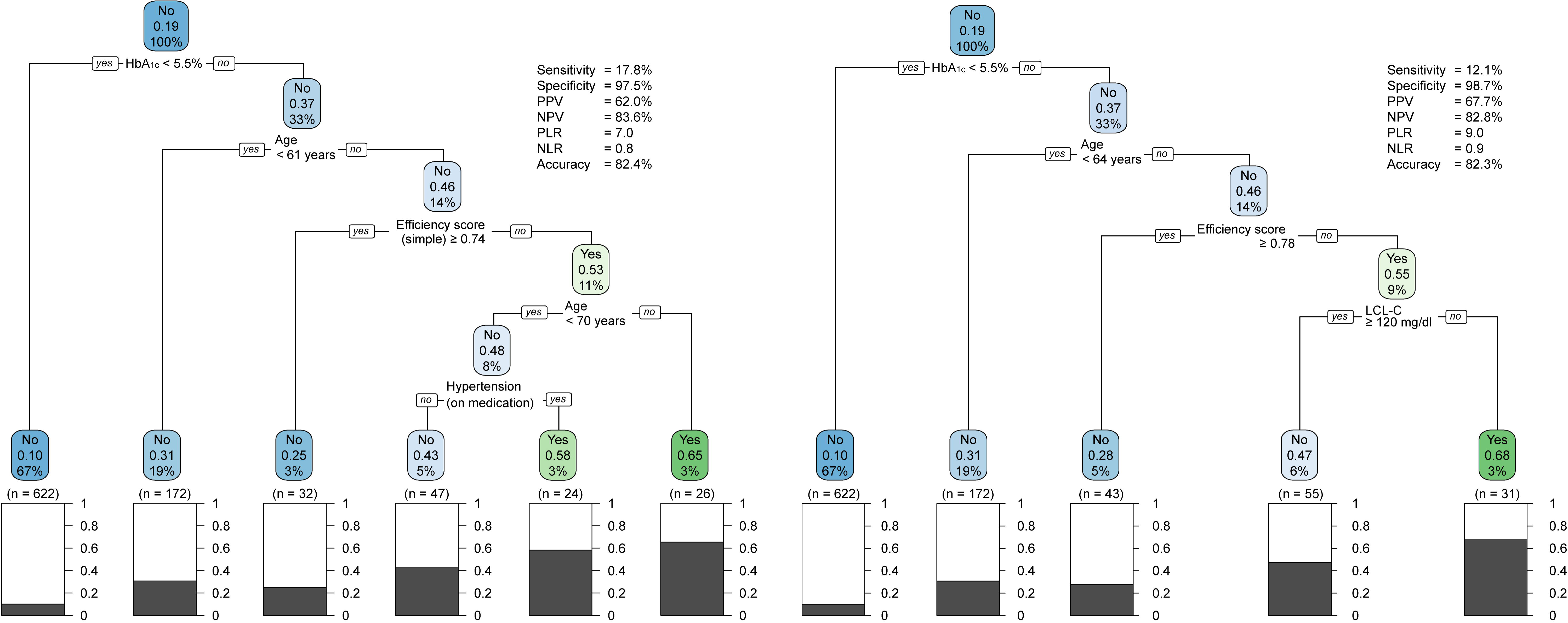
Classification tree for predicting development of prediabetes and diabetes. Regression tree on the left is the tree using efficecncy score (simple) and on the right is the tree using the efficienct score. The boxes at the bottom are the terminal nodes. The percentage shown in gray indicates participants who developed prediabetes or diabetes. For each colored box, the information on top indicates the dominant class (whether 50% or more participants developed prediabetes or diabetes) based on the percentage of participants who developed prediabetes or diabetes in the middle, and on the bottom is the percentage of total data. HbA1c, glycated hemoglobin A1c; LCL-C, low density lipoprotein cholesterol; PPV, positive predictive value; NPV, negative predictive value; PLR, positive likelihood ratio; NLR, negative likelihood ratio.

**Figure 4.**
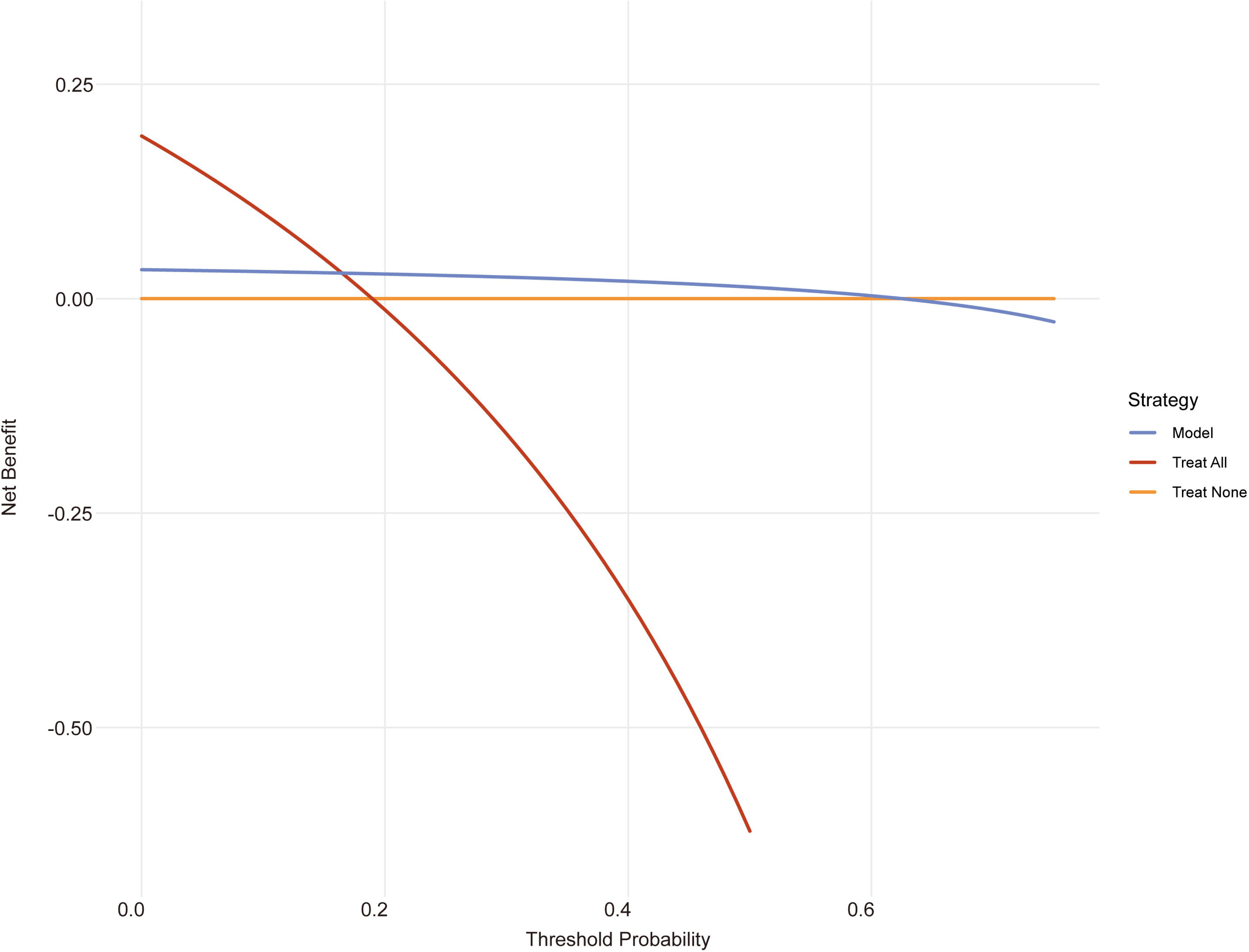
Decision curve for efficiency score model. The y-axis represents net benefit, and the x-axis represents the threshold probability. The blue line represents the Net Benefit of our model, the red line represents the ‘treat all’ strategy, and the orange line represents the ‘treat none’ strategy.

## Discussion

This study explored the hypothesis that a DEA-derived efficiency score, conceptualized as a measure of an individual’s relative efficiency in maintaining glycemic health with given physiological inputs, is associated with the future development of prediabetes among individuals with baseline HbA1c level < 5.6% at the health checkup. Our findings support this hypothesis, demonstrating that lower efficiency scores were linked to a higher risk of developing prediabetes. This suggests that even within a population screened as healthy, underlying inefficiencies in maintaining metabolic balance, as captured by the DEA score, can be indicative of future risk. By combining the efficiency score with CART analysis, we were able to identify the population likely to be prioritized for primary prevention intervention.

This study involved individuals with HbA1c values < 5.6% who were eligible for primary prevention. Approximately 85% of the participants were provided with information regarding specific health guidance, which implies that their checkup results were within the normal range (Table 1) (5). The association between the efficiency score and the risk for disease development evident in this normal population suggests that the score could serve as a useful tool for identifying the “at risk of risks” population and prioritizing them for primary prevention measures. Considering the weak correlation between the efficiency score and HbA1c levels, as well as the results from the CART analysis (Figure 3), combining these two approaches has the potential to substantially enhance the identification of the target population for primary prevention (13).

Current evidence-based approaches provide standardized interventions aimed at modifying lifestyle behaviors, including reducing calorie intake and promoting increased physical activity, to the entire population considered healthy (e.g., all 923 participants in this study) (3, 34). In contrast, our approach allows for more targeted interventions by prioritizing a subgroup of participants identified by their efficiency score and LDL-c levels (e.g., 31 participants, including 21 who developed prediabetes or diabetes in this study), we could provide more intensive interventions. Additionally, individuals in this population, who are typically notified as having “no abnormal findings” based on the conventional binary concept of healthy versus sick, often lack motivation to make necessary behavioral changes, making the promotion of healthy lifestyles challenging. Of note, the efficiency score provides a clear and easily interpretable single value indicating risk level, which can enhance individuals’ motivation. This study demonstrated the feasibility of predicting the future development of prediabetes in healthy individuals, which in turn could more effectively enable primary prevention measures.

Our strategy for predicting the future development of prediabetes by combining DEA and CART analysis demonstrated relatively high specificity and negative predictive value (NPV). While the sensitivity of our CART model was low (12.1%), this finding should be interpreted within the specific context of primary prevention among individuals who have already screened negative by conventional measures. In such scenarios, the primary goal shifts from identifying all potential future cases (high sensitivity) to precisely pinpointing a smaller, high-risk subgroup that would benefit most from targeted, potentially intensive, and resource-consuming preventing interventions. The high specificity (98.7%) is crucial in this regard, as it minimizes the number of individuals incorrectly labeled as high-risk, thereby avoiding unnecessary anxiety, follow-up, and inefficient allocation of limited healthcare resources. This trade-off–lower sensitivity for higher specificity–is often a practical consideration in primary prevention for apparently healthy populations. Our model aims to identify a manageable number of “at risk of risks” individuals who, despite normal initial screening, warrant closer attention or more intensive lifestyle counseling, which might be unfeasible if applied to a larger group identified by a more sensitive but less specific model. The clinical implication is a more focused and potentially cost-effective approach to primary prevention in this specific population. While the costs of intensive primary prevention interventions can be prohibitive (35), our approach of visualizing a healthy population “at risk of risks” can help justify allocating resources to this population, enabling more intensive interventions. Moreover, our approach uses biological examination data to identify vulnerable populations, in contrast to previous methods that relied primarily on social and economic metrics to define at-risk groups. Traditional approaches based on social determinants often inadvertently placed undue responsibility for poor health outcomes on vulnerable populations. By using biological factors to identify at-risk individuals, our method provides a more objective basis for risk assessment, potentially reducing the stigmatization of specific social or economic groups while still acknowledging the broader impact of social determinants on health (36, 37).

Our decision curve analysis results indicate that the efficiency score model is useful across a wide range of clinical scenarios (33). The model’s use is particularly recommended for threshold probabilities between 0.2 and 0.4, corresponding to situations where the harm of a false negative is approximately 1.5 to 4 times that of a false positive. In other words, our approach showed better net benefit in scenarios where missing the opportunity to intervene in 153/174 (87.9%) individuals who developed prediabetes within 3 years is considered 1.5 to 4 times less harmful than unnecessarily intervening in 10/749 (1.3%) individuals who did not develop prediabetes. Of note, the current situation corresponds to ‘treat none’ strategy, which is preferred for threshold probabilities over 0.6, where the harm of a false positive is approximately 1.5 times that of a false negative.

This study’s strength lies not only in demonstrating the efficiency score’s predictive ability but also in providing an explicit framework for its clinical implementation, combining it with classification analysis for risk stratification. DEA and CART analysis are non-parametric methods that do not require any assumptions or prerequisites such as data distribution, potentially making them advantageous for diverse populations (18, 38). This characteristic of DEA allows for population-specific optimization through relative evaluation within a given production possibility set, which may enhance its adaptability across different groups. In contrast, the evidence for the secondary prevention of diabetes was obtained through parametric analysis, which can limit the generalizability of results beyond the validated population (12, 13). The non-parametric evaluation of relative efficiency in DEA appears effective for primary prevention, as observed in studies of other conditions like hypertension across various populations (15, 39). Although we did not directly compare DEA with other predictive approaches in this study, its potential adaptability to different populations offers an intriguing avenue for future research. While further studies are needed, the combination of DEA-derived efficiency scores with CART analysis shows promise for primary prevention in diverse populations. Future comparative studies could validate this approach and explore its effectiveness across various groups.

It is important to acknowledge that DEA calculates a relative efficiency score, meaning an individual’s score is dependent on the performance of others in the specific sample. This differs from risk scores derived from regression models that quantify absolute risk based on established, weighted risk factors. Therefore, the DEA scores from this study should not be interpreted as absolute measures of prediabetes risk but rather as indicators of relative standing within this cohort of individuals with normal glucose levels. This relativity means that scores may not be directly comparable across different populations or studies without re-calibration or further validation. Furthermore, while DEA allows for flexible weighting of inputs and outputs, which can be an advantage in capturing complex relationships without a priori assumptions, it can also make the direct clinical interpretation of how specific inputs contribute to the score for an individual DMU less straightforward compared to traditional regression coefficients. Our study uses the overall efficiency score as a composite risk indicator rather than focusing on individual input weights.

One limitation of this study was that only HbA1c data were available for the baseline assessment of blood glucose levels. The FPG and OFTT are generally more effective than HbA1c in evaluating insulin resistance and secretion ability in a single test (14, 40). Some subjects with HbA1c levels of < 5.7% may have impaired fasting glucose or glucose tolerance. However, in real-world settings, such as specific health checkups in Japan, HbA1c is often the sole measure of blood glucose levels (5). The use of DEA and a classification tree in this study provided a method to compensate for the current inadequacy in evaluating glucose tolerance. Insulin resistance and insulin secretion ability are theoretically reflected in the efficiency scores of each DMU. Blood lipid status was also considered in the subsequent tree classification procedure. Another limitation was that we could only report the association between the efficiency score and future development of prediabetes. Whether earlier interventions for the at-risk population would decrease the incidence of diabetes or provide potential benefits for preventing future complications must be clarified in future prospective studies such as an RCT (41). Moreover, the study’s single-city setting in Japan and relatively small sample size (923 participants) limit its generalizability to other populations. While our sample size of 923 participants and 3-year follow-up period allowed us to detect significant associations, we acknowledge that these are limitations of our study. Future studies with larger cohorts in diverse geographical and ethnic settings and longer follow-up periods are needed to validate these findings and ensure broader applicability of the approach. Additionally, our study used data from 2008 to 2013, which may not reflect current health trends, especially considering changes that have occurred since the COVID-19 pandemic. While our findings may not capture recent shifts in risk factors or lifestyle changes, the methodological approach we’ve developed remains valid and could be applied to more recent datasets as they become available.

### Conclusions

The application of DEA to evaluate glucose tolerance showed promising potential, with the efficiency score demonstrating an association with prediabetes risk among healthy individuals. Moreover, the framework combining DEA with CART analysis offers practical insights for primary prevention in healthcare settings.

## Data Availability

The datasets generated and analyzed during the current study are not publicly available for ethical reasons, but will be available upon approval of the protocols, including the purpose of data sharing (e.g., verification of reproducibility to prevent research misconduct), by the Kaminoyama City. We cannot provide the datasets for the purpose of secondary use in research, as consent for such use has not been obtained.

## Acknowledgements

We thank Kaminoyama city for their support of this study. We would like to thank Editage for the English language editing.

## References

1. American Diabetes Association Professional Practice Committee. 2. Diagnosis and Classification of Diabetes: Standards of Care in Diabetes-2024. Diabetes Care 2024;47:S20–S42.

2. International Diabetes Federation. IDF Diabetes Atlas, 10th edition. Brussels: International Diabetes Federation; 2021. Available from: https://www.ncbi.nlm.nih.gov/books/NBK581934/.

3. American Diabetes Association Professional Practice Committee. 3. Prevention or Delay of Diabetes and Associated Comorbidities: Standards of Care in Diabetes-2024. Diabetes Care 2024;47:S43–S51.

4. Chan JC, Malik V, Jia W, et al. Diabetes in Asia: epidemiology, risk factors, and pathophysiology. JAMA 2009;301:2129–40.

5. OECD. OECD Reviews of Public Health: Japan: A Healthier Tomorrow. Paris: OECD Publishing; 2019. Available from: 10.1787/9789264311602-en.

6. Olson DE, Rhee MK, Herrick K, et al. Screening for diabetes and prediabetes with proposed A1C-based diagnostic criteria. Diabetes Care 2010;33:2184–9.

7. Heikes KE, Eddy DM, Arondekar B, et al. Diabetes Risk Calculator: a simple tool for detecting undiagnosed diabetes and prediabetes. Diabetes Care 2008;31:1040–5.

8. Hardie EA, Critchley CR, Moore SM. Prediabetes Subtypes: Patterns of Risk, Vulnerabilities, and Intervention Needs. Aust Psychol 2015;50:455–63.

9. Poltavskiy E, Kim DJ, Bang H. Comparison of screening scores for diabetes and prediabetes. Diabetes Res Clin Pract 2016;118:146–53.

10. Fujiati, II, Damanik HA, Bachtiar A, et al. Development and validation of prediabetes risk score for predicting prediabetes among Indonesian adults in primary care: Cross-sectional diagnostic study. Interv Med Appl Sci 2017;9:76–85.

11. Rajput R, Garg K, Rajput M. Prediabetes Risk Evaluation Scoring System [PRESS]: A simplified scoring system for detecting undiagnosed Prediabetes. Prim Care Diabetes 2019;13:11–5.

12. Abbasi A, Peelen LM, Corpeleijn E, et al. Prediction models for risk of developing type 2 diabetes: systematic literature search and independent external validation study. BMJ 2012;345:e5900.

13. Kengne AP, Beulens JW, Peelen LM, et al. Non-invasive risk scores for prediction of type 2 diabetes (EPIC-InterAct): a validation of existing models. Lancet Diabetes Endocrinol 2014;2:19–29.

14. Heianza Y, Hara S, Arase Y, et al. HbA1c 5.7-6.4% and impaired fasting plasma glucose for diagnosis of prediabetes and risk of progression to diabetes in Japan (TOPICS 3): a longitudinal cohort study. Lancet 2011;378:147–55.

15. Nakamura S, Narimatsu H, Nakata Y, et al. Efficiency score from data envelopment analysis can predict the future onset of hypertension and dyslipidemia: A cohort study. Sci Rep 2019;9:16309.

16. Nakamura S, Kanda S, Endo H, et al. Effectiveness of a targeted primary preventive intervention in a high-risk group identified using an efficiency score from data envelopment analysis: a randomised controlled trial of local residents in Japan. BMJ Open 2023;13:e070187.

17. Kalia NK, Miller LG, Nasir K, et al. Visualizing coronary calcium is associated with improvements in adherence to statin therapy. Atherosclerosis 2006;185:394–9.

18. Breiman L. Classification and regression trees. Boca Raton: CRC Pres; 2017.

19. Ruan Y, Lin YF, Feng YA, et al. Improving polygenic prediction in ancestrally diverse populations. Nat Genet 2022;54:573–80.

20. Ministry of Health, Labour, and Welfare. Chapter 3: Social Security from a Growth Perspective. Annual Health, Labour and Welfare Report 2017. Tokyo: Ministry of Health, Labour and Welfare; 2017.

21. Seino Y, Nanjo K, Tajima N, et al. Report of the Committee on the Classification and Diagnostic Criteria of Diabetes Mellitus (Revision for International Harmonization of HbA1c in Japan). J Jpn Diabetes Soc 2012;55:485–504.

22. The Japan Diabetes Society. Treatment Guide for Diabetes 2022-2023. Tokyo: Bunkodou; 2022. [In Japanese]

23. Charnes A, Cooper WW, Rhodes E. Measuring the efficiency of decision making units. Eur J Oper Res 1978;2:429–44.

24. Thanassoulis E PM, Despic O. Data envelopment analysis: the mathematical programming approach to efficiency analysis. The measurement of productive efficiency and productivity growth. New York: Oxford University Press; 2008.

25. Bogetoft P, Otto L. Benchmarking with DEA, SFA, and R. New York: Springer; 2011.

26. R Core Team. R: A language and environment for statistical computing. R Foundation for Statistical Computing; 2023.

27. Therneau T, Grambsch P. Modeling Survival Data: Extending The Cox Model. New York: Springer; 2000.

28. Kassambara A KM, Biecek P. survminer: Drawing Survival Curves using ‘ggplot2’. 2021. Available from: https://CRAN.R-project.org/package=survminer

29. Xue Y, Schifano ED. Diagnostics for the Cox model. Commun Stat Appl Meth 2017;24:583–604.

30. Collins GS, Reitsma JB, Altman DG, et al. Transparent Reporting of a multivariable prediction model for Individual Prognosis or Diagnosis (TRIPOD): the TRIPOD statement. Diabetic Med 2015;32:146–54.

31. De Felice F, Crocetti D, Parisi M, et al. Decision tree algorithm in locally advanced rectal cancer: an example of over-interpretation and misuse of a machine learning approach. J Cancer Res Clin Oncol 2020;146:761–5.

32. Devjak R, Burnik PT, Verdenik I, et al. Embryo quality predictive models based on cumulus cells gene expression. Balkan J Med Genet 2016;19:5–12.

33. Vickers AJ, Cronin AM, Gonen M. A simple decision analytic solution to the comparison of two binary diagnostic tests. Stat Med 2013;32:1865–76.

34. Knowler WC, Barrett-Connor E, Fowler SE, et al. Reduction in the incidence of type 2 diabetes with lifestyle intervention or metformin. N Engl J Med 2002;346:393–403.

35. Takahashi K, Nakamura S, Watanabe K, et al. Availability of Financial and Medical Resources for Screening Providers and Its Impact on Cancer Screening Uptake and Intervention Programs. Int J Environ Res Public Health 2022;19:11477.

36. Frohlich KL, Potvin L. Transcending the known in public health practice: the inequality paradox: the population approach and vulnerable populations. Am J Public Health 2008;98:216–21.

37. McLaren L, McIntyre L, Kirkpatrick S. Rose’s population strategy of prevention need not increase social inequalities in health. Int J Epidemiol 2010;39:372–7.

38. Emrouznejad A, Podinovski VV, Thanassoulis E. Data envelopment analysis: theory and applications. J Oper Res Soc 2009;60:1467–8.

39. Yu Y, Chen Y, Wang Y, et al. Is the Efficiency Score an Indicator for Incident Hypertension in the Community Population of Western China? Int J Environ Res Public Health 2021;18:10132.

40. Lorenzo C, Wagenknecht LE, Hanley AJ, et al. A1C between 5.7 and 6.4% as a marker for identifying prediabetes, insulin sensitivity and secretion, and cardiovascular risk factors: the Insulin Resistance Atherosclerosis Study (IRAS). Diabetes Care 2010;33:2104–9.

41. Inzucchi SE. Clinical practice. Diagnosis of diabetes. N Engl J Med 2012;367:542–50.

